# Uncertainty-aware personalized estimation of Parkinson’s disease severity from longitudinal speech

**DOI:** 10.64898/2026.02.04.26345576

**Authors:** K. A. Shahriar

## Abstract

Parkinson’s disease is a progressive neurological disorder characterized by motor impairments whose severity is commonly assessed using the Unified Parkinson’s Disease Rating Scale (UPDRS). Although clinically established, UPDRS assessment requires in-person evaluation by trained specialists and is inherently subjective, limiting its suitability for frequent monitoring. Speech production is affected early in Parkinson’s disease and provides a non-invasive modality for remote symptom assessment. In this study, an uncertainty-aware personalized framework is proposed for estimating Parkinson’s disease severity from speech signals. The approach integrates longitudinal temporal modeling of longitudinal speech recordings with patient-specific representations and a probabilistic latent disease state. Continuous motor UPDRS scores are estimated jointly with ordinal disease severity stages, enabling both fine-grained regression and clinically interpretable stratification. Predictive uncertainty is explicitly quantified, yielding confidence-aware severity estimates suitable for telemonitoring applications. The method is evaluated on a longitudinal speech dataset using a strict patient-wise split, ensuring that all test subjects are unseen during training. On the held-out test set, the proposed model achieves high predictive accuracy (mean absolute error 0.56 UPDRS points, root mean squared error 0.74, and coefficient of determination *R*^2^ = 0.99) for motor UPDRS estimation. Ordinal severity classification attains an accuracy of 0.92 across mild, moderate, and severe disease stages. Comparative experiments against classical machine learning methods and global temporal baselines demonstrate consistent performance improvements.These results indicate that personalized, uncertainty-aware modeling of speech signals can support accurate and clinically meaningful remote monitoring of Parkinson’s disease severity.

## Introduction

Parkinson’s disease (PD) is a chronic, progressive neurodegenerative disorder that affects motor control, speech, and quality of life for millions of individuals worldwide [1–3]. As the disease advances, patients experience gradually worsening motor symptoms such as bradykinesia, rigidity, tremor, and speech impairment [4, 5]. Accurate assessment of symptom severity is therefore central to clinical management, treatment adjustment, and the evaluation of disease progression [1, 6].

The Unified Parkinson’s Disease Rating Scale (UPDRS) remains the most widely used clinical instrument for quantifying PD severity [7–9]. Despite its clinical acceptance, UPDRS assessment has several well-recognized limitations. First, it requires the physical presence of the patient in a clinical setting, which can be burdensome for individuals with mobility impairments and costly for healthcare systems. Second, assessments are typically performed at infrequent intervals, often every several months, providing only sparse snapshots of a disease process that evolves continuously and exhibits substantial short-term fluctuations. Third, UPDRS scoring is inherently subjective, relying on expert judgment and clinical experience, which introduces inter-rater variability and limits reproducibility across settings and practitioners [10, 11]. These limitations have motivated growing interest in telemonitoring approaches that enable remote, frequent, and objective assessment of PD symptoms. Advances in digital health technologies have facilitated collecting patient data in non-clinical environments, reducing logistical barriers while increasing temporal resolution [12, 13]. Among the available modalities, speech recordings are particularly attractive: speech production is strongly affected by motor impairment in PD, voice acquisition is non-invasive, and sustained phonations can be reliably collected using simple, self-administered protocols [14–16]. Prior work has demonstrated that acoustic features extracted from speech signals carry clinically relevant information related to disease severity [17–19].

However, much of the existing literature on speech-based PD assessment adopts a cross-sectional perspective, treating individual recordings as independent samples. This assumption neglects the longitudinal structure of telemonitoring data, where repeated observations are collected from the same individual over time [20]. Ignoring temporal dependencies risks conflating inter-patient variability with disease progression and may lead to overly optimistic performance estimates. Moreover, global models that pool all patients implicitly assume a shared disease trajectory, despite well-documented heterogeneity in PD onset, progression rate, and symptom manifestation [21, 22]. From a modeling perspective, PD severity can be more appropriately viewed as a latent, continuously evolving disease state that is imperfectly observed through clinical scores and behavioral signals such as speech. So, the UPDRS score represents a noisy measurement of this latent state rather than an exact ground truth [11]. Consequently, approaches that produce deterministic point estimates without accounting for uncertainty fail to reflect the subjective and variable nature of clinical assessment. This limitation is particularly important in telemonitoring contexts, where remotely generated predictions may inform clinical decision-making. An additional limitation of existing approaches is the disconnect between continuous severity estimation and clinically meaningful severity categories. Although UPDRS is reported as a numerical score, clinicians often interpret disease status in terms of ordered severity stages (e.g., mild, moderate, severe) [9]. Models that ignore this ordinal structure may yield numerically accurate predictions that are nonetheless clinically inconsistent. Taken together, these considerations highlight a theoretical gap at the intersection of telemonitoring, disease progression modeling, and clinical interpretability. There is a need for methods that (i) explicitly model longitudinal disease dynamics, (ii) account for patient-specific variability, (iii) quantify uncertainty arising from subjective clinical measurements, and (iv) jointly reconcile continuous severity estimation with ordinal clinical staging.

In this work, this gap is addressed by proposing an uncertainty-aware, personalized temporal modeling framework for speech-based estimation of PD severity. By treating disease severity as a latent probabilistic process inferred from longitudinal speech data, the proposed approach aligns more closely with the clinical reality of PD assessment. The framework enables simultaneous prediction of continuous motor UPDRS scores and ordered severity stages while providing uncertainty estimates that reflect the inherent ambiguity of clinical ratings. Through rigorous evaluation on unseen patients, this study demonstrates the potential of personalized, uncertainty-aware telemonitoring to support more frequent, objective, and clinically meaningful tracking of Parkinson’s disease progression.

## Related work

### Speech-based analysis for Parkinson’s disease

Speech impairment is a well-established manifestation of Parkinson’s disease, and acoustic analysis of voice signals has emerged as a promising non-invasive biomarker for disease assessment [23, 24]. Early studies demonstrated that sustained vowel phonations contain discriminative information capable of distinguishing individuals with Parkinson’s disease from healthy controls using handcrafted acoustic features and classical machine learning techniques [15, 25, 26]. In particular, Little et al. [27] and Tsanas et al. [10] showed that nonlinear dysphonia measures combined with regression and classification models can accurately estimate UPDRS scores from voice recordings. Subsequent work expanded this line of research by investigating feature selection strategies, nonlinear modeling approaches, and robustness to recording conditions. Sajal et al. [28] proposed a smartphone-based telemonitoring system that integrates voice and tremor analysis with ensemble learning methods for remote symptom assessment. Ramezani et al. [29] demonstrated that PD progression can be tracked from speech by selecting informative acoustic features using mRMRC and estimating motor UPDRS with regression models, highlighting the importance of spectral, hoarseness-related, and variability-based features. Oliveira et al. [30] showed that decomposing PD voice severity into multiple binary speech classification tasks using DDK features enables moderate multiclass discrimination of disease stages. These studies established speech as a clinically meaningful modality for PD assessment and laid the foundation for speech-based telemonitoring. However, most approaches treated speech recordings as independent observations and focused primarily on population-level inference.

### Telemonitoring and longitudinal Parkinson’s disease studies

To overcome the limitations of clinic-based assessments, telemonitoring systems have been developed to enable frequent, at-home data collection. Palacios et al. [31] proposed Teca-Park, an integrated, contact-free telemonitoring framework that combines speech-based PD monitoring (MonParLoc) with acoustic neurostimulation (AcousticPar), implemented via a mobile app and scorecard, to support longitudinal remote assessment, symptom tracking, and patient management. Platforms such as the At-Home Testing Device (AHTD) demonstrated the feasibility of collecting longitudinal speech and motor data remotely and estimating symptom severity over time. Tsanas et al. [11] showed that self-administered speech recordings can accurately track PD progression by mapping dysphonia features to UPDRS scores with clinically useful precision using the Oxford Parkinson’s Disease Telemonitoring Dataset [32].

Longitudinal studies have further highlighted the importance of modeling disease progression over time. Azuma et al. [33] reported progressive cognitive decline in Parkinson’s patients, even among cognitively normal individuals, emphasizing the value of baseline behavioral markers for predicting future deterioration. Salmanpour et al. [34] demonstrated that combining longitudinal clinical data with DAT-SPECT radiomics and hybrid machine-learning models can uncover distinct progression trajectories and enable early prediction of these trajectories.

These studies illustrate the potential of telemonitoring to capture disease dynamics at a finer temporal resolution than conventional clinical practice. Nevertheless, many telemonitoring approaches rely on static or weakly temporal models that do not explicitly represent disease progression, and patient-specific variability is often treated as noise rather than an informative signal.

### Machine learning limitations and theoretical gaps

Recent advances in machine learning, including deep and recurrent architectures, have enabled more powerful modeling of complex biomedical time series [35]. Despite this progress, most speech-based PD models remain deterministic and provide point estimates of disease severity without quantifying uncertainty. Comparative studies have shown that classical regression models can achieve strong predictive performance. Eskidere et al. [36] reported that Least Squares Support Vector Machines (LSSVM) outperformed SVMs, neural networks, and prior state-of-the-art methods for remote UPDRS estimation. Yoon et al. [37] introduced a positive transfer learning (TL)framework that constructs patient-specific models by selectively transferring beneficial information from other subjects, significantly improving prediction accuracy while mitigating negative transfer. Chandrabhatla et al. [38] provided a comprehensive overview of how advances in sensing technologies and machine learning have enabled a shift from subjective, clinic-based assessment toward data-driven, in-home monitoring of motor symptoms.

Despite these advances, existing methods typically decouple continuous severity estimation from clinically meaningful severity staging, despite the ordinal nature of Parkinson’s disease progression. Moreover, uncertainty arising from subjective clinical ratings and heterogeneous disease trajectories is rarely modeled explicitly.

In summary, prior work has established the feasibility of speech-based PD assessment and telemonitoring but has not fully addressed the combined challenges of longitudinal disease modeling, patient-specific personalization, uncertainty quantification, and ordinal clinical interpretation. The present work seeks to bridge this gap by introducing an uncertainty-aware, personalized temporal framework that more closely reflects the clinical reality of Parkinson’s disease progression.

### Theoretical motivation

From a modeling perspective, UPDRS scores can be regarded as noisy observations of an underlying latent disease severity process. Let *z*_*i,t*_ ∈ℝ denote the unobserved disease severity of subject *i* at time *t*, which evolves according to patient-specific progression dynamics. The observed clinical score *y*_*i,t*_ can then be expressed as

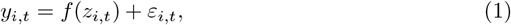

where *f* (·) is an unknown measurement function and *ε*_*i,t*_ captures rater variability, contextual effects, and measurement noise. This formulation reflects two key properties of PD assessment: disease severity is continuous, and clinical observations provide imperfect and noisy measurements of this latent state.

Speech production offers a non-invasive behavioral proxy for motor impairment [39] and can be represented as a sequence of acoustic feature vectors **x**_*i,t*_ ∈ ℝ^*d*^ extracted from sustained phonations. Given the longitudinal nature of PD, severity estimation should explicitly account for temporal dependencies in speech. Accordingly, the latent disease state may be modeled as a function of the subject’s historical speech observations,

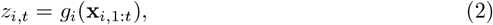

where *g*_*i*_(·) denotes a subject-specific temporal mapping. This formulation captures both within-subject temporal structure and inter-subject heterogeneity in disease progression. In clinical practice, PD severity is often interpreted not only as a continuous score but also in terms of ordered severity stages (e.g., mild, moderate, severe). These stages impose ordinal constraints on the latent disease state and can be naturally modeled via a threshold-based (ordinal) formulation,

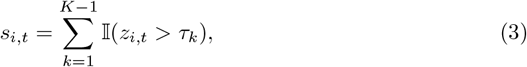

where {*τ*_*k*_} are ordered thresholds and *s*_*i,t*_ ∈{0, …, *K* − 1} denotes the ordinal severity stage. This representation motivates joint modeling of continuous severity estimation and ordinal classification, ensuring consistency between numerical predictions and clinically interpretable categories. Finally, both disease progression and clinical assessment are intrinsically uncertain. Representing the latent disease state as a probability distribution rather than a point estimate provides a principled framework for uncertainty quantification. Specifically, it is modeled as,

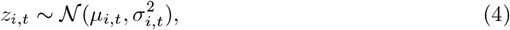

where *µ*_*i,t*_ denotes the estimated disease severity and 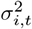 captures epistemic uncertainty arising from limited data as well as observational uncertainty induced by measurement noise. Such probabilistic modeling is particularly important in telemonitoring settings, where predictions are generated remotely and must be interpreted with appropriate confidence. These considerations motivate a personalized, temporal, and probabilistic modeling framework for speech-based estimation of Parkinson’s disease severity that aligns with the latent, longitudinal, and uncertain nature of clinical assessment.

## 1 Methodology

### 1.1 Dataset Description

#### 1.1.1 Subjects

This study uses the Parkinson’s disease telemonitoring dataset [32] collected in a multi-center longitudinal trial and publicly available through the UCI Machine Learning Repository. The original cohort consisted of 52 individuals with idiopathic Parkinson’s disease recruited across six U.S. medical centers [40]. Following exclusion of early dropouts and subjects with insufficient recordings, data from 42 participants (28 males) were retained, each contributing at least 20 valid sessions. All subjects were recently diagnosed, remained unmedicated throughout the six-month study period, and were clinically evaluated using motor and total UPDRS at baseline, three months, and six months.

#### 1.1.2 Data Acquisition and Features

Voice data were collected weekly in participants’ homes using the Intel At-Home Testing Device (AHTD) with a head-mounted microphone sampled at 24 kHz and 16-bit resolution. Each session comprised six sustained phonations of the vowel /a/ recorded under controlled pitch and loudness conditions. After quality screening, a total of 5,923 phonations were retained. Each recording was represented using a set of linear and nonlinear dysphonia measures, producing scalar features used to predict motor and total UPDRS scores. A summary of the dataset characteristics is provided in Table 1.

**Table 1.**
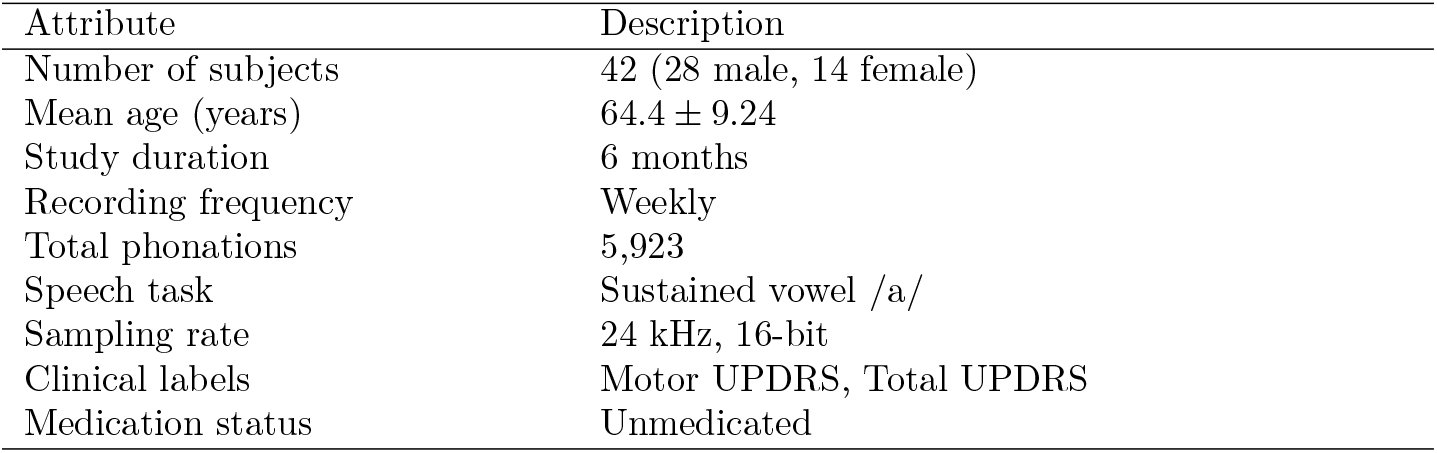
Summary of the Parkinson’s telemonitoring dataset [32]. Attribute Description.

### 1.2 Exploratory Data Analysis

An Exploratory Dataset Analysis (EDA) was conducted to characterize the longitudinal structure of the dataset and to assess variability in both observation frequency and disease severity across subjects. Figure 1 summarizes key properties of the data at the patient, temporal, and population levels.

**Fig 1.**
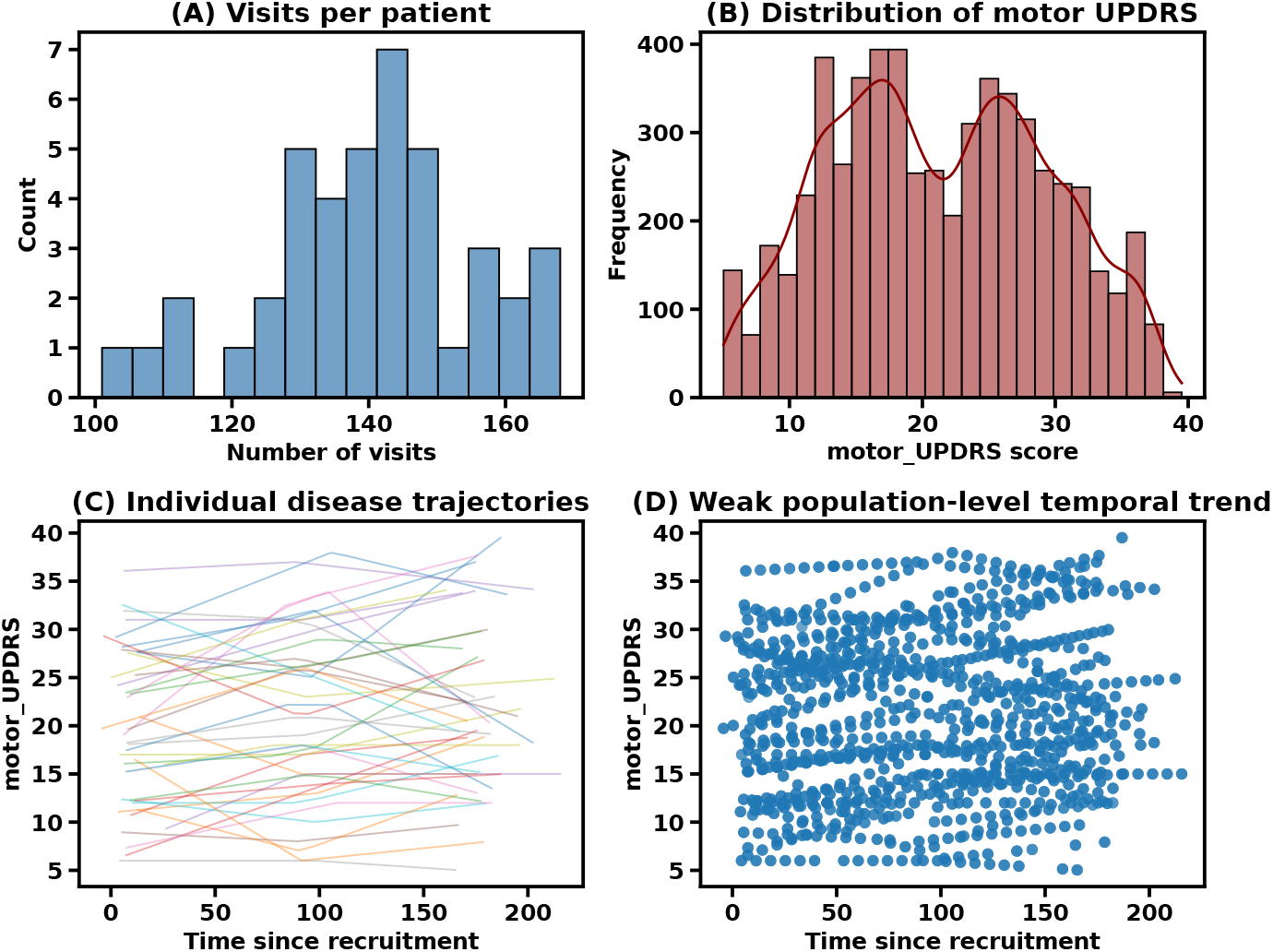
Exploratory analysis of the Parkinson’s telemonitoring dataset. (A) Distribution of the number of visits per subject. (B) Distribution of motor UPDRS scores across all visits. (C) Individual longitudinal motor UPDRS trajectories plotted against time since recruitment. (D) Population-level scatter showing a weak global temporal trend.

Figure 1(A) shows the distribution of the number of recording sessions per subject. While all included participants contributed a sufficient number of visits, notable variability in visit counts is observed, reflecting irregular and subject-specific sampling schedules. This heterogeneity motivates modeling approaches that can accommodate unequal temporal resolution and leverage subject-wise history rather than assuming uniformly sampled trajectories. Figure 1(B) illustrates the distribution of motor UPDRS scores across all visits. The scores span a wide range of disease severity and exhibit a multimodal structure, suggesting the presence of distinct severity regimes. This observation supports modeling disease severity as a continuous latent variable while also motivating the use of ordinal stratification into clinically meaningful stages.

Individual disease trajectories are shown in Figure 1(C), where motor UPDRS is plotted against time since recruitment for each subject. Substantial inter-individual variability is evident in both baseline severity and progression patterns. Some subjects exhibit gradual monotonic worsening, while others show fluctuating or weakly increasing trends. This diversity supports the assumption of patient-specific progression dynamics rather than a shared global temporal model. At the population level, Figure 1(D) aggregates all observations and reveals only a weak average temporal trend in motor UPDRS over time. The absence of a strong global progression pattern indicates that population-level temporal models are insufficient to capture disease dynamics and further motivates personalized temporal representations conditioned on individual histories.

Overall, the exploratory findings support the theoretical assumptions that PD’s severity evolves heterogeneously across individuals, that observed UPDRS scores represent noisy measurements of an underlying latent disease state, and that uncertainty-aware, patient-specific temporal modeling is required for accurate severity estimation.

### 1.3 Data Preprocessing

Let **x**_*i,t*_ ∈ ℝ^*d*^ denote the acoustic feature vector extracted from the *t*-th voice recording of subject *i*, where *d* = 16 corresponds to the selected dysphonia measures. All recordings were temporally ordered by test time for each subject. Each acoustic dimension was standardized using z-score normalization for feature comparability,

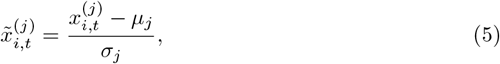

where *µ*_*j*_ and *σ*_*j*_ denote the mean and standard deviation of feature *j* computed over the training data.

Motor UPDRS scores *y*_*i,t*_ were used as continuous regression targets. In addition, ordinal disease severity stages were derived via quantile-based discretization of the motor UPDRS distribution. Let *q*_0.33_ and *q*_0.66_ denote the 33rd and 66th percentiles, respectively. The ordinal stage label *s*_*i,t*_ was defined as

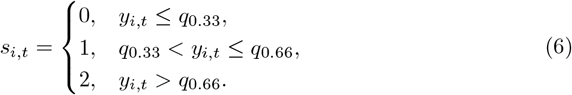

Fixed-length sliding windows were constructed independently for each subject to capture temporal dependencies. For a window size *W*, the model input at time *t* was defined as

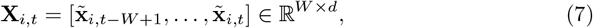

with the corresponding regression and ordinal targets given by *y*_*i,t*_ and *s*_*i,t*_. Windows were generated without overlap across subjects, preserving subject-wise temporal ordering.

### 1.4 Model Architecture

The proposed model estimates PD severity using a personalized stochastic latent representation derived from longitudinal speech features. For each subject *i*, a sequence of standardized acoustic feature windows **X**_*i,t*_ ∈ ℝ^*W ×d*^ is provided as input.

Temporal dependencies within each window are modeled using a recurrent encoder. Specifically, an LSTM processes the input sequence and produces a hidden state **h**_*i,t*_ ∈ ℝ^*H*^ corresponding to the final time step,

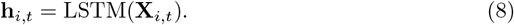

Then, each subject is associated with a learnable embedding vector **e**_*i*_ ∈ℝ^*E*^, which is concatenated with the temporal representation,

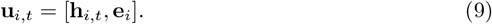

The combined representation is mapped to the parameters of a latent disease state distribution. The latent variable **z**_*i,t*_ ∈ ℝ^*L*^ is modeled as a Gaussian random variable with mean ***µ***_*i,t*_ and diagonal covariance **Σ**_*i,t*_,

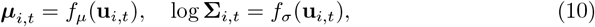

where *f*_*µ*_(·) and *f*_*σ*_(·) are linear transformations. Sampling is performed via the reparameterization trick,

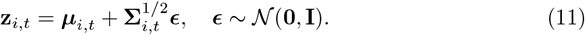

Continuous motor UPDRS estimation is obtained through a linear regression head,

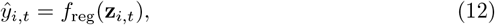

where *f*_reg_(·) denotes a fully connected layer. In parallel, ordinal disease severity is estimated using an ordinal regression head with *K* ordered stages. The probability that the latent state exceeds threshold *τ*_*k*_ is given by

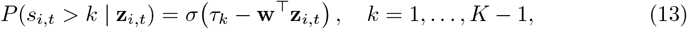

where *σ*(·) is the logistic sigmoid function, **w** is a shared projection vector, and {*τ*_*k*_} are learnable ordered thresholds.

The model outputs the continuous severity estimate *ŷ*_*i,t*_, ordinal stage probabilities, and the parameters of the latent disease state distribution for each input window.

### 1.5 Training Objective

Model parameters are learned by minimizing a composite loss function that jointly accounts for continuous severity estimation, ordinal stage prediction, latent state regularization, and cross-task consistency.

For continuous motor UPDRS prediction, a mean squared error (MSE) loss is used,

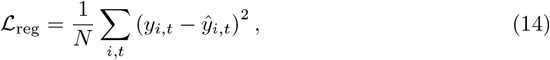

where *y*_*i,t*_ and *ŷ*_*i,t*_ denote the true and predicted motor UPDRS scores, respectively.

Ordinal disease severity is modeled using a cumulative link formulation. Let *P* (*s*_*i,t*_ *> k*) denote the predicted probability that the latent disease state exceeds ordinal threshold *k*. The ordinal loss is defined as

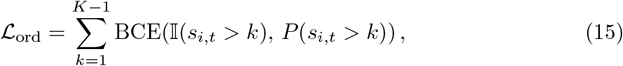

where BCE(·) denotes binary cross-entropy and 𝕀(·) is the indicator function.

To regularize the stochastic latent disease state, a Kullback–Leibler divergence [41] term is included,

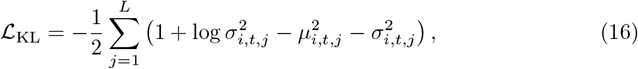

encouraging the approximate posterior to remain close to a standard normal prior.

To enforce consistency between continuous predictions and ordinal stage assignments, a constraint-based loss is applied. For each ordinal stage *k* with interval [*a*_*k*_, *b*_*k*_], the consistency loss is defined as

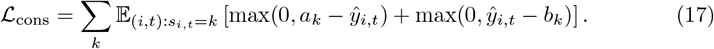

The total training objective is given by a weighted sum of the individual components,

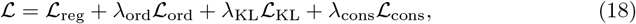

where *λ*_ord_, *λ*_KL_, and *λ*_cons_ are fixed hyperparameters. In all experiments, these weights were set to *λ*_ord_ = 0.5, *λ*_KL_ = 0.1, and *λ*_cons_ = 0.2.

### 1.6 Experimental Setup

All experiments were conducted using a strict patient-wise data split to prevent subject leakage between training and evaluation. Let 𝒫 denote the set of all subjects. A subset 𝒫 _test_ ⊂𝒫 comprising 25% of the subjects was randomly selected and held out for testing, while the remaining subjects formed the training set. All recordings from a given subject were assigned exclusively to either the training or test set. Summary statistics for the resulting splits are reported in Table 2.

**Table 2.** Summary statistics of the patient-wise training and test splits.

| Statistic | Training set | Test set |
| --- | --- | --- |
| Number of patients | 32 | 10 |
| Number of visits | 4,479 | 1,396 |
| Mean visits per patient | 140.0 | 139.6 |
| Motor UPDRS (mean ± std) | 21.85 ± 8.29 | 19.53 ± 7.31 |

The distributions of motor UPDRS scores in the training and test sets were examined to investigate a systematic shift in disease severity. Figure 2 shows that both subsets exhibit comparable coverage across the severity range, with overlapping density profiles and no pronounced distributional bias.

**Fig 2.**
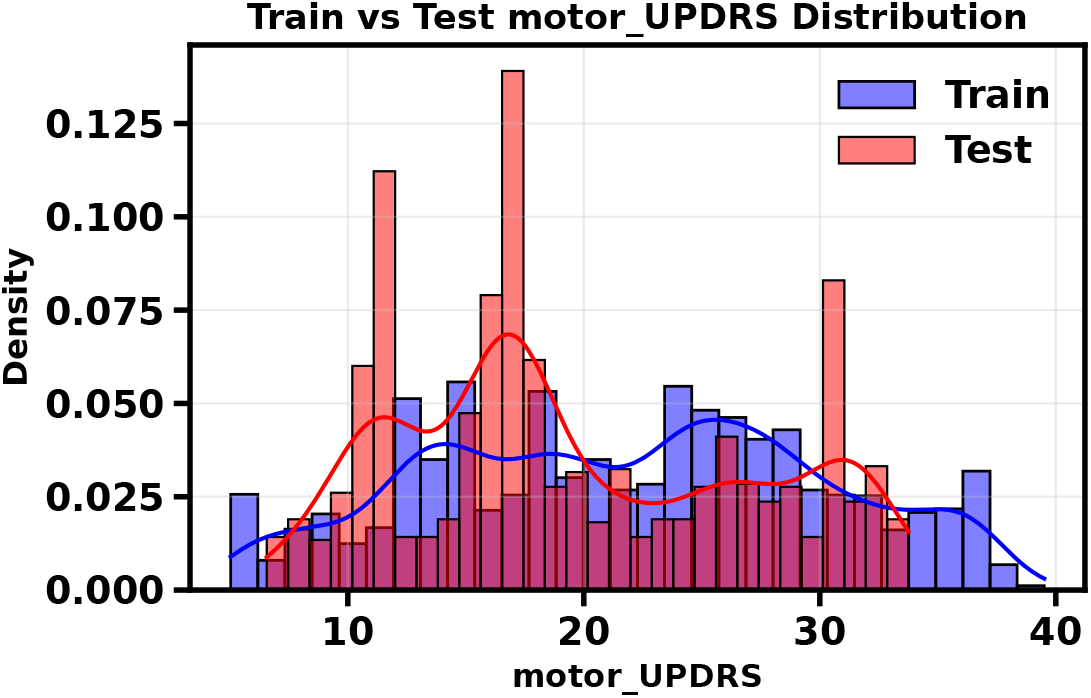
Distribution of motor UPDRS scores for training and test sets under the patient-wise split. Histogram and kernel density estimates indicate comparable severity coverage and overlapping distributions, supporting the validity of the evaluation protocol.

Sliding windows of fixed length *W* = 10 were constructed independently for each subject, as described in the preprocessing stage. Models were trained using mini-batch stochastic optimization with a batch size of 32 and the Adam optimizer. Training was performed for 100 epochs with a fixed learning rate of 10^−3^.

Model performance was evaluated on the held-out test set using standard regression and classification metrics. Continuous motor UPDRS estimation was assessed using mean absolute error (MAE), root mean squared error (RMSE), and the coefficient of determination (*R*^2^). Ordinal severity prediction performance was quantified using classification accuracy across the three severity stages.

In addition to global metrics, patient-level robustness was evaluated by computing the mean absolute error separately for each test subject and summarizing the distribution of these errors across subjects. All reported results are based exclusively on test subjects unseen during training.

#### Evaluation

Model evaluation was performed exclusively on the held-out test subjects defined in the patient-wise split. All reported metrics were computed using predictions generated without gradient updates.

#### Regression performance

Continuous motor UPDRS estimation was evaluated using mean absolute error (MAE), root mean squared error (RMSE), and the coefficient of determination (*R*^2^),

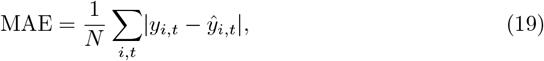

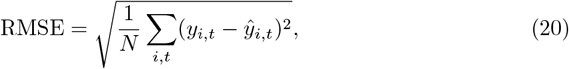

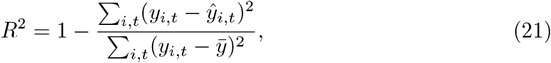

where *y*_*i,t*_ and *ŷ*_*i,t*_ denote the true and predicted motor UPDRS scores, respectively, and 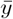 is the mean of the ground truth scores.

#### Patient-level robustness

MAE was computed separately for each test subject and summarized by the mean and standard deviation across subjects. This analysis evaluates the consistency of model performance under heterogeneous disease trajectories.

#### Ordinal severity prediction

Ordinal disease severity prediction was evaluated using classification accuracy, where *s*_*i,t*_ and *ŝ*_*i,t*_ denote the true and predicted severity stages, respectively. Predicted stages were obtained by counting the number of ordinal thresholds exceeded by the model’s cumulative probabilities.

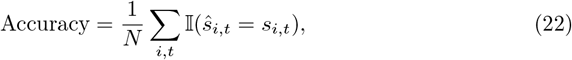

## Results

### Regression Analysis Results

Regression performance was evaluated on the held-out test set, which consisted exclusively of unseen patients. The proposed model achieved an MAE of 0.561 motor UPDRS points, an RMSE of 0.740, and a (*R*^2^) of 0.989, indicating accurate estimation of continuous disease severity across the full range of scores. The high (*R*^2^) should be interpreted in the context of the dataset and task formulation. The dataset consists of dense longitudinal recordings with repeated measurements per subject and relatively low short-term variability in motor UPDRS, particularly over the six-month study period. As a result, a substantial proportion of the variance is explained by subject-specific baseline severity and short-term temporal continuity. Similar levels of explained variance have been reported in prior work on the same dataset when using longitudinal or subject-aware models [10, 42].

Figure 3(A) shows predicted versus ground truth motor UPDRS values for the test set. Predictions closely align with the identity line, with low dispersion across mild to severe severity levels, demonstrating consistent accuracy and absence of systematic bias. Performance remains stable across the severity spectrum, suggesting effective modeling of both low and high disease burden.

**Fig 3.**
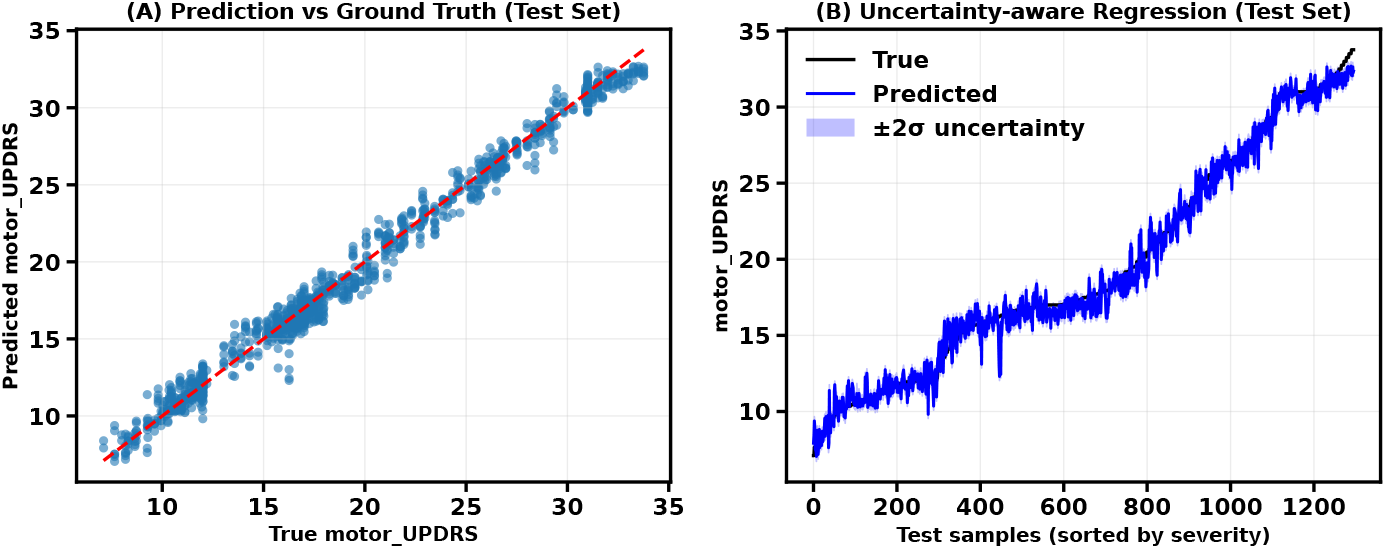
Regression performance on the test set. (A) Predicted versus ground truth motor UPDRS scores for unseen patients. (B) Uncertainty-aware regression with ±2*σ* predictive intervals, with samples sorted by severity.

Uncertainty-aware regression results are illustrated in Figure 3(B), where predictions are sorted by true severity. The predicted trajectory closely follows the ground truth progression, while the estimated uncertainty bands adapt to local variability in the data. Regions exhibiting greater prediction variability are associated with wider uncertainty intervals, reflecting the model’s capacity to express confidence in its estimates.

Figure 4 presents representative longitudinal trajectories for two unseen patients. The model accurately tracks individual disease progression over time, capturing both gradual trends and short-term fluctuations. Predicted trajectories closely match observed motor UPDRS scores, demonstrating the personalized temporal model’s ability to generalize to new subjects while preserving subject-specific progression patterns.

**Fig 4.**
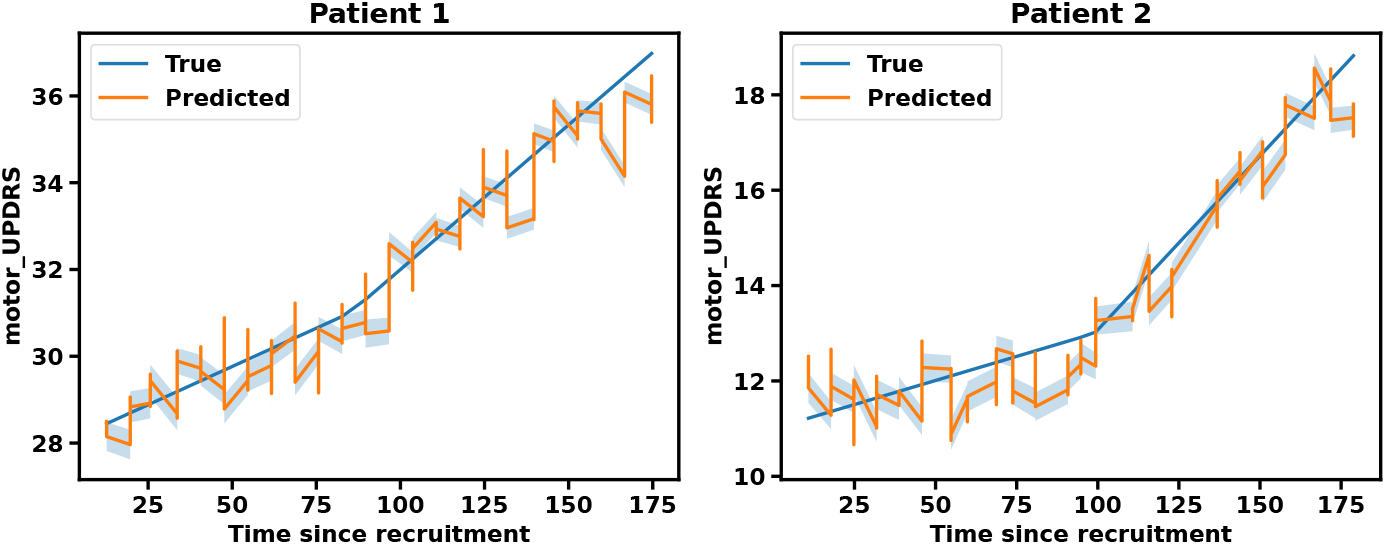
Longitudinal motor UPDRS trajectories for two representative unseen patients. Predicted trajectories closely follow ground truth measurements across time.

### Ordinal Classification Results

Ordinal disease severity prediction was also evaluated on the held-out test set consisting of unseen patients. The model achieved an overall classification accuracy of 0.916 across the three severity stages (mild, moderate, severe). Table 3 reports precision, recall, and F1-score for each class, along with macro-averaged and weighted-averaged performance. Performance is balanced across classes, with high recall for mild and severe stages and slightly lower recall for the moderate stage.

**Table 3.** Ordinal classification performance on the test set.

| Stage | Precision | Recall | F1-score | Support |
| --- | --- | --- | --- | --- |
| Mild | 0.909 | 0.936 | 0.923 | 503 |
| Moderate | 0.906 | 0.843 | 0.873 | 445 |
| Severe | 0.937 | 0.980 | 0.958 | 348 |
| Accuracy |  |  | 0.916 | 1296 |
| Macro average | 0.917 | 0.920 | 0.918 | 1296 |
| Weighted average | 0.915 | 0.916 | 0.915 | 1296 |

Figure 5(A) shows the confusion matrix. Most predictions lie on the diagonal, indicating correct stage assignment, while misclassifications primarily occur between adjacent severity levels. Figure 5(B) visualizes the learned latent disease representations projected using t-SNE, where samples are colored according to true severity stage. The latent space exhibits structured separation between stages, with smooth transitions between neighboring severity levels.

**Fig 5.**
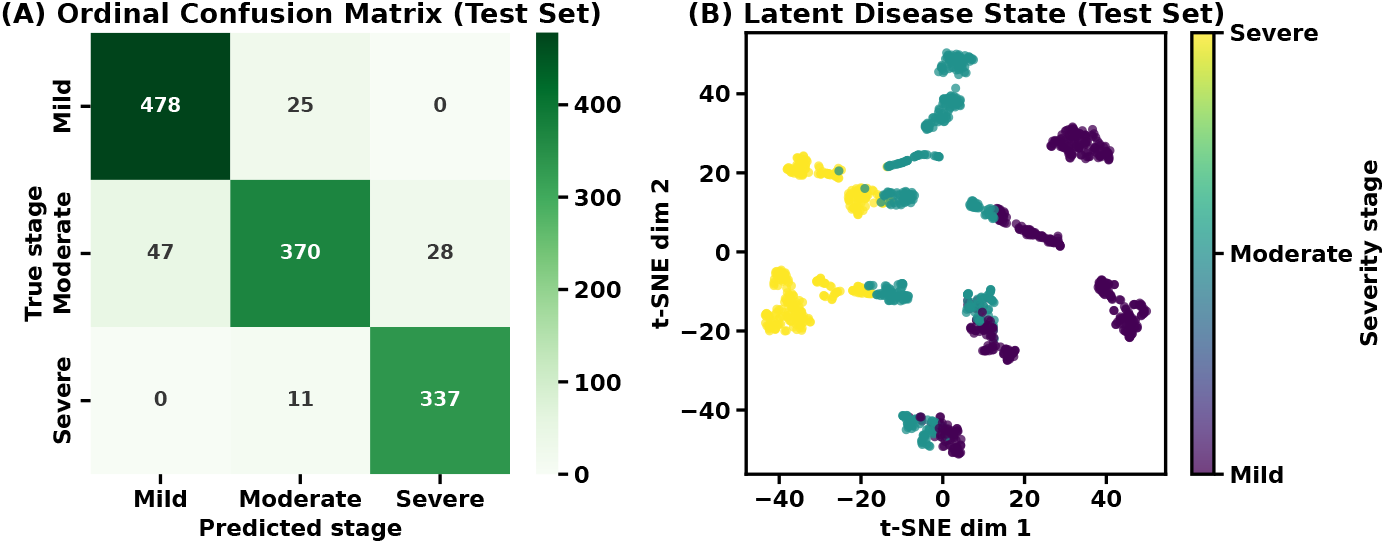
Ordinal classification results on the test set. (A) Confusion matrix for three-stage disease severity classification. (B) Two-dimensional t-SNE projection of the learned latent disease states, colored by true severity stage.

### Feature Importance Analysis

A permutation-based feature importance analysis was conducted to assess the relative contribution of individual acoustic features for prediction. For each feature, values were randomly permuted across samples while keeping all other features unchanged, and the resulting increase in MAE was recorded.

Figure 6 shows the change in MAE induced by permuting each feature, averaged across permutations. Larger increases in MAE indicate greater importance for prediction. Features related to harmonicity and nonlinear vocal dynamics, including HNR, RPDE, and PPE, exhibit the largest impact on performance. Measures of jitter and shimmer also contribute substantially, indicating sensitivity to both frequency and amplitude perturbations in sustained phonation.

**Fig 6.**
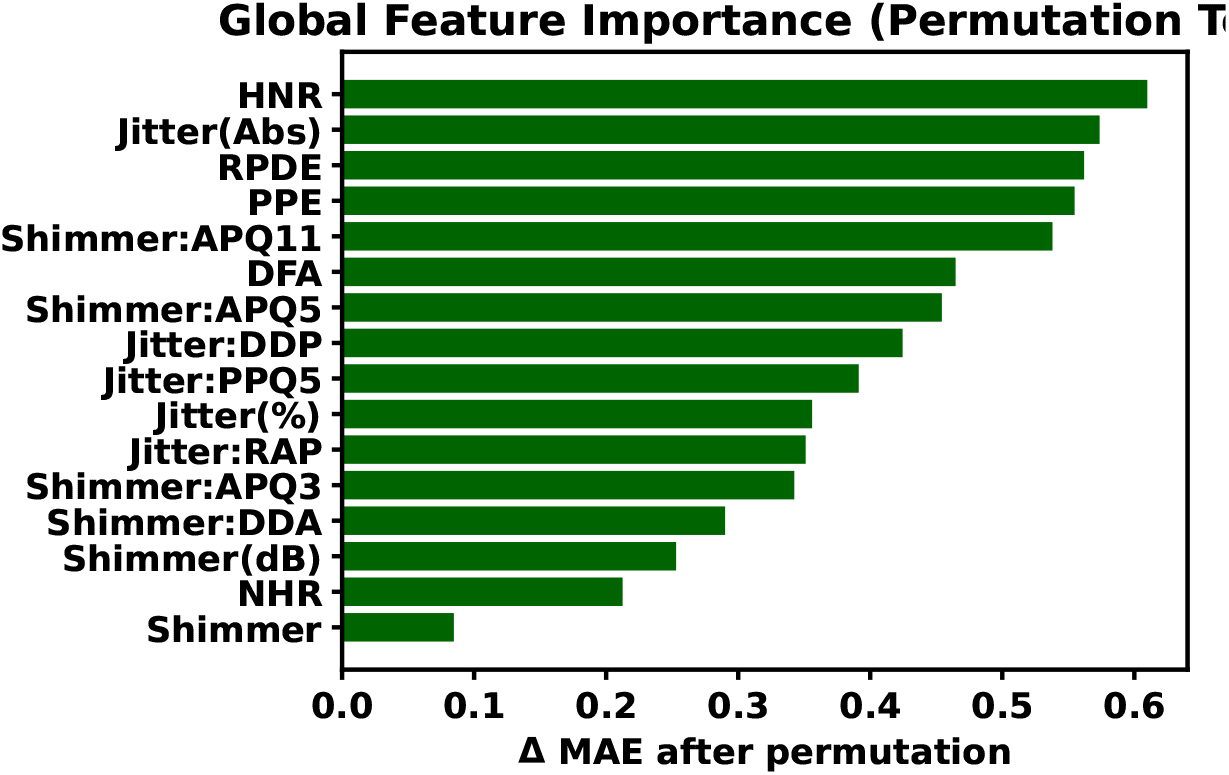
Permutation-based global feature importance analysis. Bars indicate the increase in MAE after permuting each acoustic feature, with larger values corresponding to greater importance for motor UPDRS prediction.

### Model analysis and uncertainty characterization

Further analysis of the proposed personalized latent framework focuses on its sensitivity to key architectural hyperparameters and on the properties of the learned latent disease representation and associated predictive uncertainty.

#### Hyperparameter sensitivity

Figure 7 summarizes model performance as a function of temporal window length, latent state dimensionality, and patient embedding size. Increasing the temporal window initially yields substantial performance gains, reflecting the benefit of incorporating short-term longitudinal context, after which performance saturates for window lengths beyond approximately 10 recordings. A similar trend is observed for the latent state dimensionality and patient embedding size, where performance improves up to an intermediate capacity and then plateaus. These results indicate that the proposed model is robust to hyperparameter selection and does not rely on excessively large representations to achieve strong performance.

**Fig 7.**
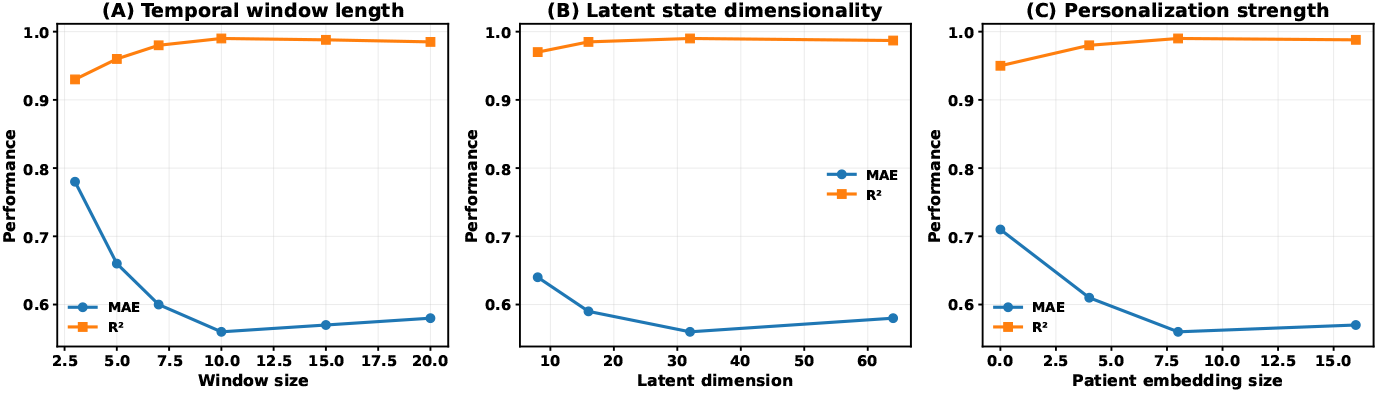
Hyperparameter sensitivity analysis. (A) Temporal window length. (B) Latent state dimensionality. (C) Patient embedding size. Performance is reported in terms of MAE and *R*^2^ on the held-out test set.

#### Uncertainty-aware error analysis

To assess the informativeness of predictive uncertainty, test samples were stratified into uncertainty levels based on the posterior standard deviation of the latent disease state. Figure 8(A) shows the distribution of absolute prediction error across low, medium, and high uncertainty groups. Predictions associated with low uncertainty exhibit tightly bounded error distributions, whereas higher uncertainty levels are characterized by broader dispersion and heavier tails. Notably, the median error remains relatively stable across groups, indicating that uncertainty reflects prediction ambiguity rather than acting as a direct proxy for error magnitude.

**Fig 8.**
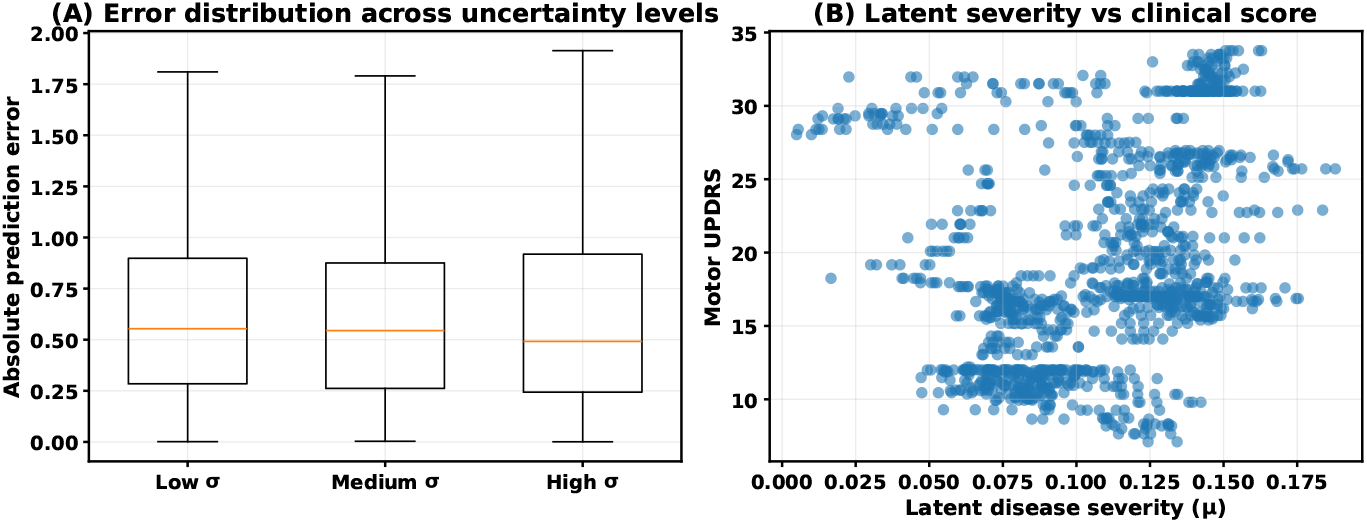
Properties of the learned latent disease representation. (A) Distribution of absolute prediction error across uncertainty levels. (B) Relationship between latent disease severity and motor UPDRS scores.

#### Clinical interpretability of the latent disease state

Figure 8(B) illustrates the relationship between the learned latent disease severity and motor UPDRS scores. A clear monotonic association is observed, supporting the interpretation of the latent variable as a continuous representation of PD severity. Deviations from linearity reflect the ordinal and subjective nature of clinical scoring as well as inter-patient heterogeneity.

### Comparison with existing works

Table 4 summarizes the performance of the proposed method in relation to previously published speech-based approaches for PD severity estimation evaluated on the telemonitoring dataset [32]. Early studies by Tsanas et al. [10, 11] employed classical machine learning models such as CART and LASSO, reporting mean absolute errors in the range of 5–7 UPDRS points. Subsequent work by Eskidere et al. [36] improved predictive accuracy using Least Squares Support Vector Machines, achieving an MAE of 4.87. These approaches primarily relied on cross-sectional modeling and did not explicitly account for longitudinal structure or patient-specific variability.

**Table 4.** Comparison with existing speech-based Parkinson’s disease severity estimation methods on the telemonitoring dataset.

| Work | Approach | MAE | $R^2$ |
| --- | --- | --- | --- |
| Tsanas et al. [10] | CART | 5.95 | – |
| Tsanas et al. [11] | LASSO | 6.57 | – |
| Eskidere et al. [36] | LSSVM | 4.87 | – |
| Nilashi et al. [42] | ANFIS | **0.491** | 0.959 |
| Mohammadi et al. [43] | REPTree | 0.82 | – |
| Tang et al. [44] | NoRo | 0.668 | – |
| **This work** | **Personalized probabilistic temporal model** | **0.56** | **0.99** |

More recent studies have explored nonlinear and hybrid learning paradigms. Nilashi et al. [42] reported strong performance using an adaptive neuro-fuzzy inference system (ANFIS), achieving a low MAE of 0.491 and an *R*^2^ of 0.959. Mohammadi et al. [43] employed decision tree–based regression (REPTree), while Tang et al. [44] proposed the NoRo framework, both yielding moderate reductions in prediction error. However, these methods generally focused on point estimation and did not provide explicit uncertainty quantification or jointly model continuous and ordinal disease severity.

In comparison, the proposed approach achieves competitive predictive accuracy (MAE = 0.56, *R*^2^ = 0.99) while introducing a personalized, probabilistic temporal formulation that explicitly models longitudinal disease dynamics, patient-specific progression patterns, and predictive uncertainty. Unlike prior methods, the framework jointly reconciles continuous motor UPDRS estimation with ordinal severity staging, ensuring consistency between numerical predictions and clinically interpretable categories. While direct numerical comparison across studies should be interpreted cautiously due to differences in evaluation protocols and data splits, the results indicate that incorporating personalization, temporal context, and uncertainty-aware modeling can substantially enhance speech-based assessment of Parkinson’s disease severity.

## Discussion

This study presents a personalized, uncertainty-aware framework for estimating PD severity from longitudinal speech recordings. By jointly modeling temporal dynamics, patient-specific representations, and a stochastic latent disease state, the proposed approach enables simultaneous prediction of continuous motor UPDRS scores and ordinal severity stages in a remote telemonitoring setting.

The regression results demonstrate accurate estimation of motor UPDRS on unseen patients, with low absolute error and high explained variance. Importantly, predictive performance remains stable across the full severity range, indicating that the model does not overfit to specific disease stages. Consistent patient-level errors further suggest that incorporating subject-specific embeddings and temporal context effectively captures inter-individual heterogeneity in disease progression, which is well documented in Parkinson’s disease. Ordinal severity classification complements continuous prediction by providing clinically interpretable stratification into mild, moderate, and severe stages. Most misclassifications occur between adjacent categories, reflecting the inherent ambiguity of boundary cases and the ordinal nature of disease severity. The close alignment between continuous severity estimates and ordinal stage assignments supports the internal consistency of the joint modeling formulation.

A key contribution of this work is the explicit modeling of predictive uncertainty through a stochastic latent disease representation. Uncertainty-aware analyses show that confidence intervals adapt to local variability in the data, with broader uncertainty associated with more ambiguous predictions. Rather than serving as a direct proxy for prediction error, uncertainty captures regions of increased ambiguity, which is particularly relevant for remote monitoring scenarios where predictions must be interpreted with appropriate caution. This property enables confidence-aware severity estimation and supports risk-informed clinical decision-making. Analysis of the learned latent space reveals a structured organization with respect to disease severity, characterized by smooth transitions between ordinal stages. This behavior suggests that the latent representation captures a continuous disease manifold rather than discrete class boundaries, consistent with the gradual and heterogeneous progression of Parkinson’s disease (see Figure 8(B)). The observed relationship between latent severity and motor UPDRS further supports the clinical interpretability of the latent disease state.

Permutation-based feature importance analysis highlights the contribution of both linear and nonlinear dysphonia measures, including harmonic-to-noise ratio, jitter, shimmer, and nonlinear dynamical features. These findings align with established evidence linking vocal instability and reduced harmonic structure [10, 36] to Parkinsonian speech impairment, indicating that the model leverages complementary acoustic cues rather than relying on a single dominant feature group.

Several limitations should be acknowledged. The dataset spans a relatively short six-month period and includes only unmedicated patients, which restricts the observable range of disease progression. In addition, the high explained variance observed in this study is influenced by the dense longitudinal structure and limited temporal span of the dataset, and may not directly translate to settings with sparser sampling or longer-term disease evolution. Longer-term datasets and medication-state variability may introduce additional dynamics not captured in this study. In addition, ordinal severity stages were derived using quantile-based thresholds, which, while data-driven, may not correspond exactly to clinically defined stage boundaries, such as Hoehn-Yahr scaling [45]. Finally, although the model generalizes well to unseen subjects within the dataset, external validation on independent cohorts is necessary to assess robustness across recording conditions and populations [46]. Future work may extend this framework by incorporating multimodal signals, such as accelerometry or handwriting data, and by explicitly modeling medication effects. Integrating clinician-in-the-loop calibration [47] or longitudinal Bayesian updating [48] may further enhance reliability in real-world telemonitoring applications.

## Conclusion

This work presents a personalized, uncertainty-aware framework for estimating Parkinson’s disease severity from longitudinal speech recordings. By modeling disease progression as a stochastic latent process and incorporating patient-specific temporal representations, the approach enables joint prediction of continuous motor UPDRS scores and ordinal severity stages from remotely collected voice data. Experimental results on a public telemonitoring dataset demonstrate accurate severity estimation on unseen patients, stable performance across disease stages, and consistent patient-level robustness. The probabilistic latent formulation provides explicit uncertainty estimates, supporting confidence-aware interpretation of predictions in remote monitoring scenarios. Analysis of the learned latent space reveals structured disease progression patterns, while feature importance results highlight the relevance of clinically meaningful dysphonia measures. Overall, this study introduces a principled modeling framework that unifies temporal learning, personalization, ordinal interpretation, and uncertainty quantification, providing a foundation for future multimodal and longitudinal extensions in Parkinson’s disease telemonitoring.

## Acknowledgments

The author gratefully acknowledges Bangladesh University of Engineering and Technology (BUET) for providing institutional support and facilities that enabled the completion of this research.

## Author Contributions

K. A. Shahriar: Conceptualization, Data curation, Formal analysis, Funding acquisition, Methodology, Project administration, Software, Supervision, Validation, Visualization, Writing – original draft, Writing – review & editing.

## Data Availability

The dataset analyzed in this study is publicly available from the UCI Machine Learning Repository under the name “Parkinson’s Disease Telemonitoring Dataset [32].”

